# Modelling, Simulations and Analysis of the First and Second COVID-19 Epidemics in Beijing

**DOI:** 10.1101/2021.04.02.21254821

**Authors:** Lequan Min

**Affiliations:** School of Mathematics and Physics, University of Science and Technology Beijing, Beijing, 100083, PR China

**Keywords:** New coronavirus, SIR model, Disease-free equilibrium, Globally attractive, Simulations, Long-term’s estimation

## Abstract

To date, over 130 million people on infected with COVID-19. It causes more 2.8 millions deaths. This paper introduces a symptomatic-asymptomatic-recoverer-dead differential equation model (SARDDE). It gives the conditions of the asymptotical stability on the disease-free equilibrium of SARDDE. It proposes the necessary conditions of disease spreading for the SARDDE. Based on the reported data of the first and the second COVID-19 epidemics in Beijing and simulations, it determines the parameters of SARDDE, respectively. Numerical simulations of SARDDE describe well the outcomes of current symptomatic and asymptomatic individuals, recovered symptomatic and asymptomatic individuals, and died individuals, respectively. The numerical simulations suggest that both symptomatic and asymptomatic individuals cause lesser asymptomatic spread than symptomatic spread; blocking rate of about 90% cannot prevent the spread of the COVID19 epidemic in Beijing; the strict prevention and control strategies implemented by Beijing government is not only very effective but also completely necessary. The numerical simulations suggest also that using the data from the beginning to the day after about two weeks at the turning point can estimate well or approximately the following outcomes of the two COVID-19 academics, respectively. It is expected that the research can provide better understanding, explaining, and dominating for epidemic spreads, prevention and control measures.

## 1 Introduction

In December 2019, a novel coronavirus-induced pneumonia (COVID-19) broke out in Wuhan, Hubei. Now over one hundred million people on infected with COVID-19 have been identified worldwide. It causes more 2.2 millions deaths. It affects more than 220 countries and regions including Antarctica. One of the reasons of such a tragedy is that people in some countries do not pay attention to theoretical analysis and estimations for COVID-19 epidemic. In fact mathematical models for epidemic infectious diseases have played important roles in the formulation, evaluation, and prevention of control strategies. Modelling the dynamics of spread of disease can help people to understand the mechanism of epidemic diseases, formulate and evaluate prevention and control strategies, and predict tools for the spread or disappearance of an epidemic [1].

Since the outbreak of COVID-19 in Wuhan, many scholars have published a large numbers of articles on the modeling and prediction of COVID-19 epidemic (for examples see [2–9]). It is difficult to describe well the dynamics of COVID-19 epidemics. In a Lloyd-Smith et al’s paper, it described nine challenges in modelling the emergence of novel pathogens, emphasizing the interface between models and data [10].

In January 19, two Beijing people coming back from Wuhan were confirmed to be infected with COVID-19. Thus has caused the first wave COVID-19 epidemic in Beijing. A total of 411 locally diagnosed cases were reported during the first wave COVID-19 epidemic. After 140 days, 411 COVID-19 patients were cured, and 9 patients died. The medical personnel has realized the zero infection.

This paper introduces a symptomatic-asymptomatic-recoverer-dead differential equation model (SARDDE). It gives the conditions of the asymptotical stability on the disease-free equilibrium of SARDDE. Using simulations determines the parameters of SARDDE based on the reported data of COVID-19 epidemic in Beijing [11]. Numerical simulations of SARDDE describe well the practical outcomes of current infected symptomatic and asymptomatic individuals, recovered infected symptomatic and asymptomatic individuals, and died infected individuals.

The rest of this paper is organized as follows. Section 2.1 establishes SARDDE. Section 2.2 provides the criterions of the asymptotical stability of the disease-free equilibrium. Section 2.3 determines the necessary conditions of disease spreading. Section 3.1 implements the dynamic simulations of SARDDE to describe the data of the first COVID-19 epidemic in Beijing; states analysis and discussions. In the same section two virtual simulation examples are implemented to emphasize the importance of strict control measures and long terms’ estimation to epidemic spreading. Section 3.2.1 establishes a model without died case. Section 3.2.2 provides the criterions of the asymptotical stability of the disease-free equilibrium. Section 3.2.3 determines the necessary conditions of disease spreading. Section 3.2.4 implements the dynamic simulations of the model to describe the data of the second COVID-19 epidemic in Beijing; states analysis and discussions. In the same subsection, two virtual simulation examples are implemented. Conclusions are given in Section 4.

## 2 SARDDE Model and Dynamic Properties

### 2.1 1.1 SARDDE Model

For SARDDE model, there are four states. *I*(*t*), *I*_*a*_(*t*), *I*_*r*_(*t*), *I*_*ra*_(*t*) and *D*(*t*) represent the fraction of current symptomatic infected individuals, and current asymptomatic but infected individuals, cumulative recovered symptomatic infected individuals, cumulative recovered asymptomatic but infected individuals and cumulative died individuals, respectively. The transition among these states is governed by the following rules (Flowchart of the rules is shown in Fig.1, where *S* represents susceptible population.).

**Figure 1:**
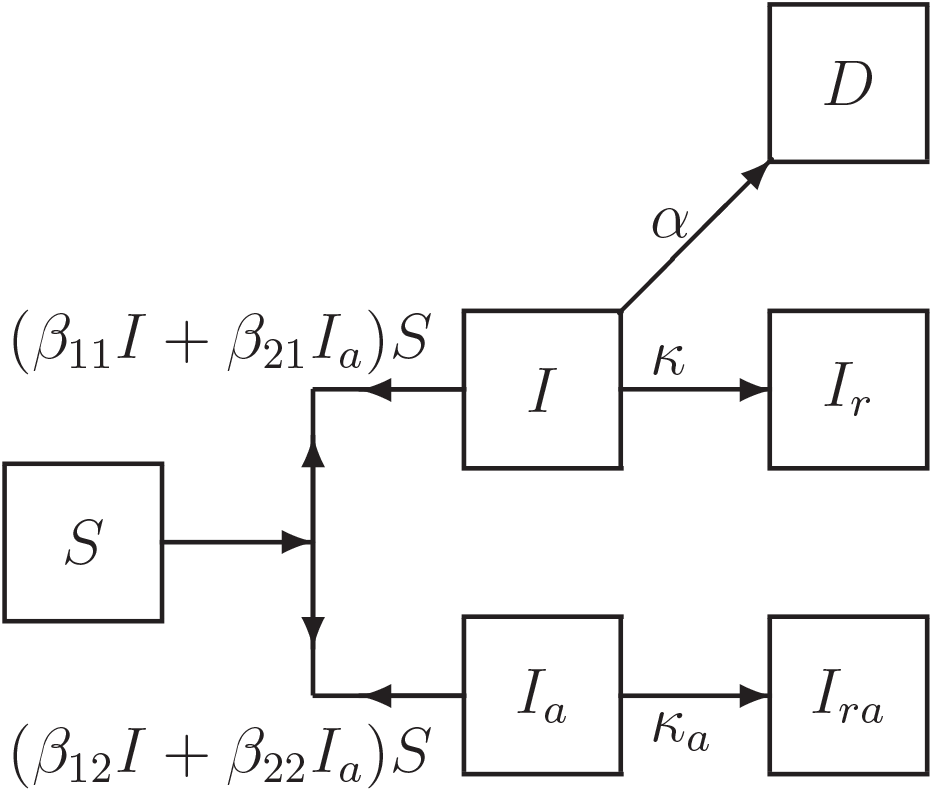
Flowchart of disease transmission among susceptible population S, current symptomatic infected individuals I, current asymptomatic but infected individuals *I*_*a*_ recovered symptomatic infected individuals *I*_*r*_, recovered asymptomatic but infected individuals *I*_*ra*_, and died individuals *D*.

First, the symptomatic infected individuals (*I*) and the asymptomatic but infected individuals (*I*_*a*_) infect the susceptible population (*S*) with the probabilities of *β*_11_ and *β*_21_, respectively, making *S* become symptomatic infected individuals, and with the probabilities of *β*_12_ and *β*_22_, respectively, making *S* become asymptomatic individuals. Then, a symptomatic individual is cured at a rate *κ*, an asymptomatic individual returns to normal at a rate *κ*_*a*_. An infected individual dies at a rate *α*. Here all parameters are positive numbers. Assume that the dynamics of an epidemic can be described by *m* time intervals. At *i*th interval, the model has the form:

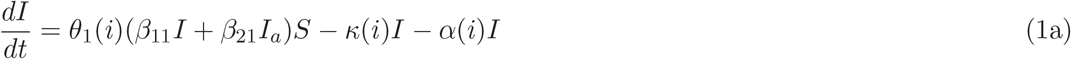

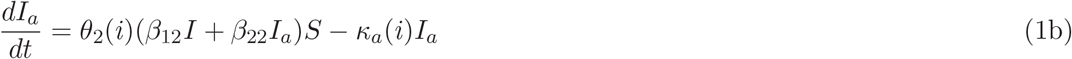

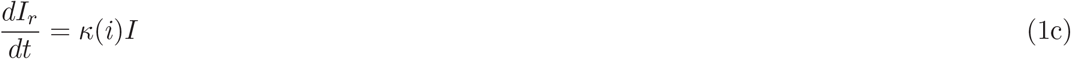

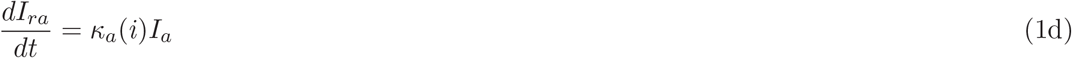

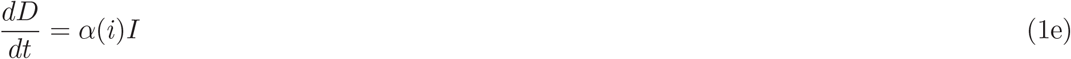

where *θ*_1_(*i*)^′^*s* and *θ*_1_(*i*)^′^*s* (*i* = 1, *…, m*) represent blocking rates to symptomatic and asymptomatic infections, respectively. Then system (1) has a disease-free equilibrium:

Then equation (1) has a disease-free equilibrium:

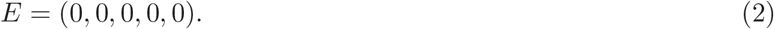

### 2.2 Stability of disease-free equilibrium

The stability of system (1) is determined by the first two equations (1a) and (1b). Denote in (1a) and (1b)

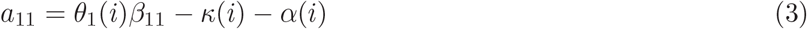

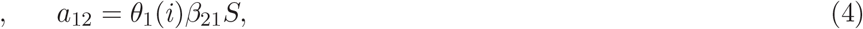

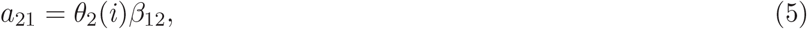

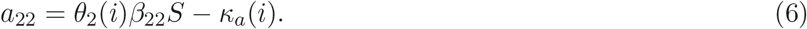

Then at the disease-free equilibrium of system(18), the Jacobian matrix of (18a) and (18b) is

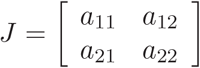

Solving the corresponding eigenequation obtains 2 eigenvalues:

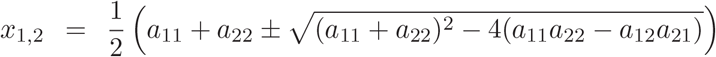

Therefore it obtains the following:

#### Theorem 1

*Suppose that a*_11_, *a*_12_, *a*_21_ *and a*_22_ *are defined by (3)-(6) then the disease-free equilibrium E of system (18) is globally asymptotically stable if, and only if, the following inequalities hold:*

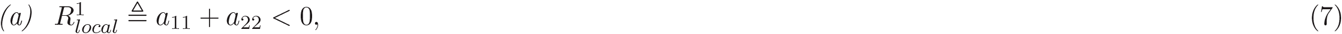

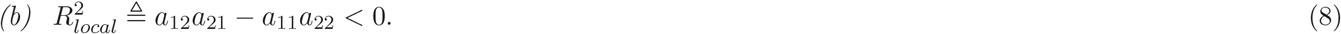

### 2.3 The necessary condition of disease spreading

If an epidemic can occur, then

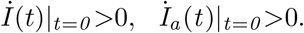

This implies that

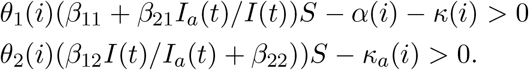

Solving the above inequalities gives the following

#### Theorem 2

*If system (1) satisfy the following inequalities*

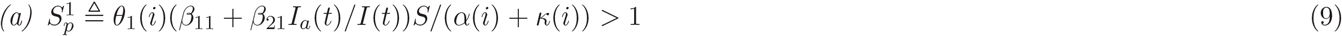

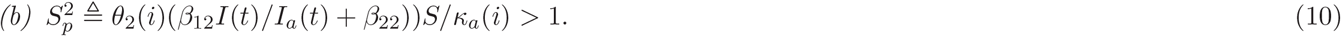

*then a disease transmission will occur*.

## 3 Applications

Based on the reported clinical COVID-19 epidemic data from January 19 to June 8, 2020 in Beijing [11], this Section will discuss the applications of the above theoretical results. Numerical simulations and drawings are performed by using MATLAB software programs. The first 50 days’ reported clinical data on current confirmed infection cases, and the reported clinical data on recovered cases of the COVID-19 epidemic in Beijing [11] are shown in Figs. 2(a) and 2(b)^1^. The number of current symptomatic infected individuals is showed in Fig3(a) by circles. The numbers of cumulative recovered symptomatic infected individuals, and cumulative died infected individuals are showed in Fig3(b) by circles and stars respectively.

**Figure 2:**
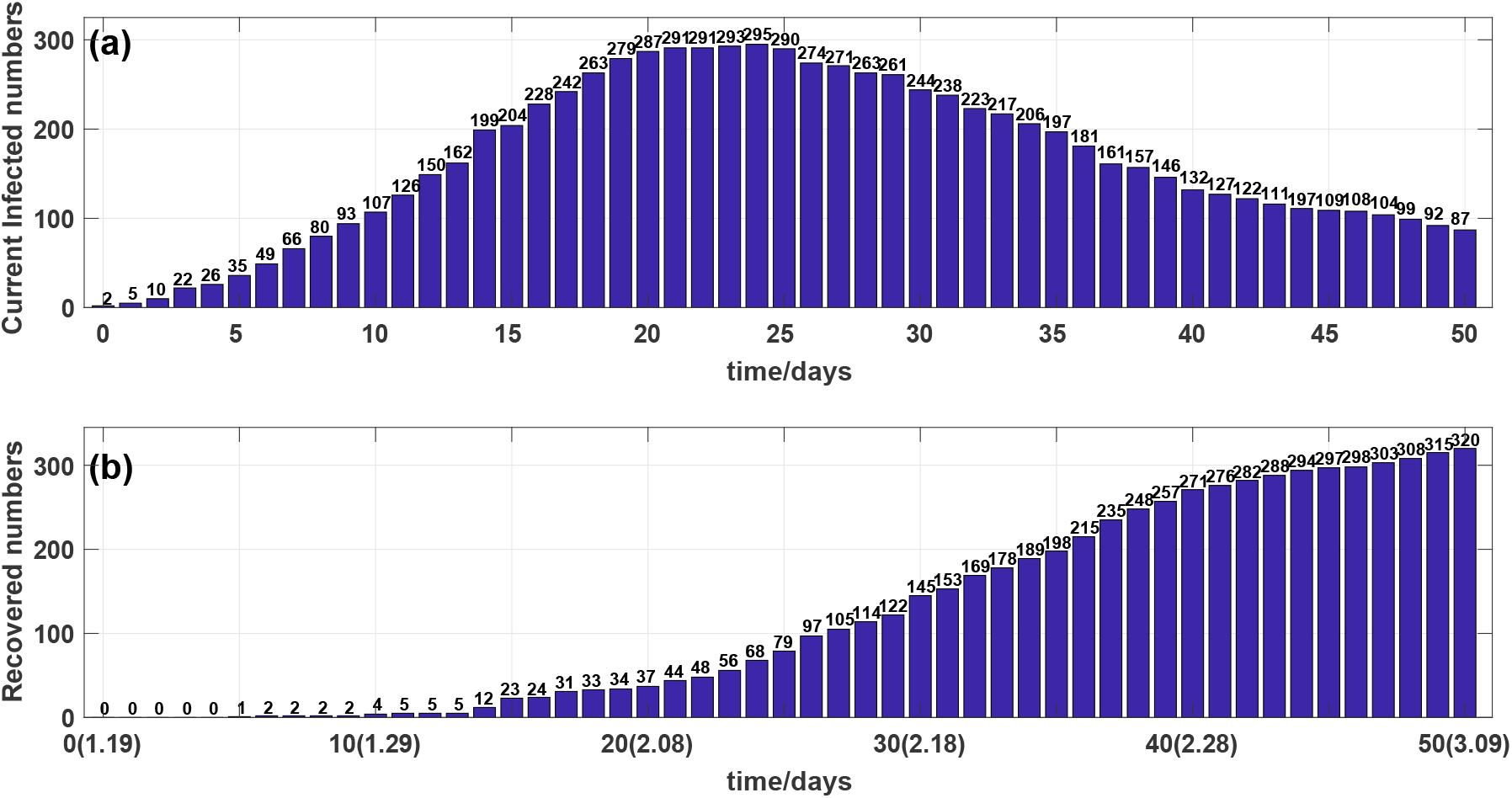
(a) Outcome of the number of current infected individuals. (b) Outcome of the number of cumulative recovered individuals.

**Figure 3:**
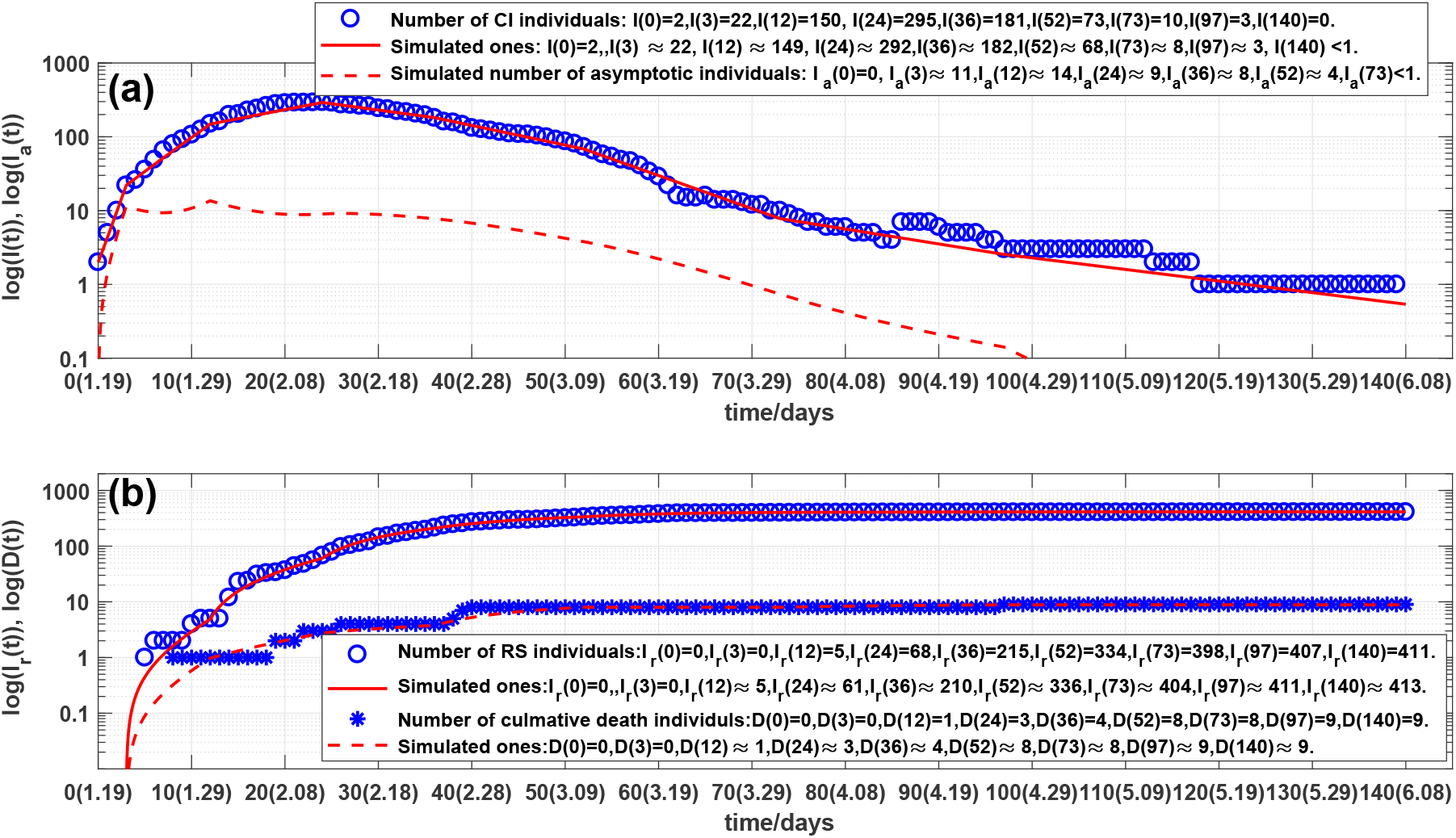
(a) Outcome of the number of current symptomatic individuals, representing by circles. Solid line and dash line are outcomes of stimulated current symptomatic individuals and stimulated current asymptomatic individuals of system (1). (b) Outcomes of the numbers of cumulative recovered symptomatic and died individuals, representing by circles and stars, respectively. Solid line and dash line are corresponding simulations of system (1).

The number of current infected individuals was risen rapidly in the first 4 days (see Fig. 2(a)). The number of current infected individuals reached the highest 295 on the day 24th, February 12 and then after the day 31th, February 19, declined rapidly (see Fig. 2(a) and 3(a)).

Observe from the Figs. 3(a) and 3(b) that the overall changes in the number of current confirmed infections are not subject to the law of exponential changes, but the data can be approximated in good agreement with 8 straight lines in log scale (see Fig. 3). This phenomenon can be explained as: different medical measures prevention and control strategies have been adopted at the different 8 time intervals. On the day 86th, April 15, there are 3 Chaoyang district infected people coming back Beijing form foreign country which makes calculated blacking rates to rise. Therefore the *i* in SARDDE model (1) should be chosen as *i* = 1, 2, *…*, 8.

### 3.1 Simulation and prediction of the first COVID-19 epidemic in Beijing

First it needs to determine the parameters *κ*(*i*), *κ*_*a*_(*i*) and *α*(*i*). There are different methods for calculating the recovery rate *κ*(*i*) in a specific time interval. Denote *s*_1_(*i*) and *s*_2_(*i*) to be the days that the old patients and the new patients stayed in the hospital during *i*th time interval. Denote *R*(*i*) and *d*(*i*) to be the numbers of the recovered patients and died patients during *i*th time interval, respectively. Similar to the formula given in Ref. [12], *R*(*i*) and *d*(*i*) can be defined by

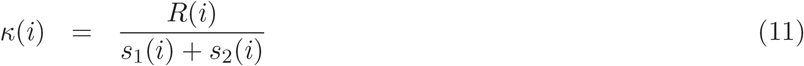

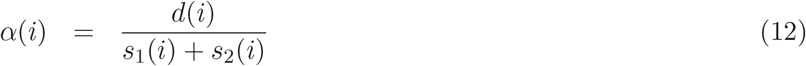

Since there is no information on recovered asymptomatic infected individuals, we take

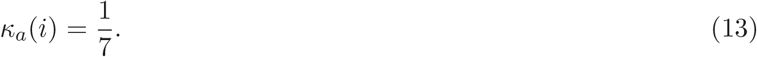

That is, an asymptomatic infected individual will recover in average 7 days. The calculated *κ*(*i*)^′^*s* and *α*(*i*)^′^*s* are shown in Table 1.

**Table 1.**
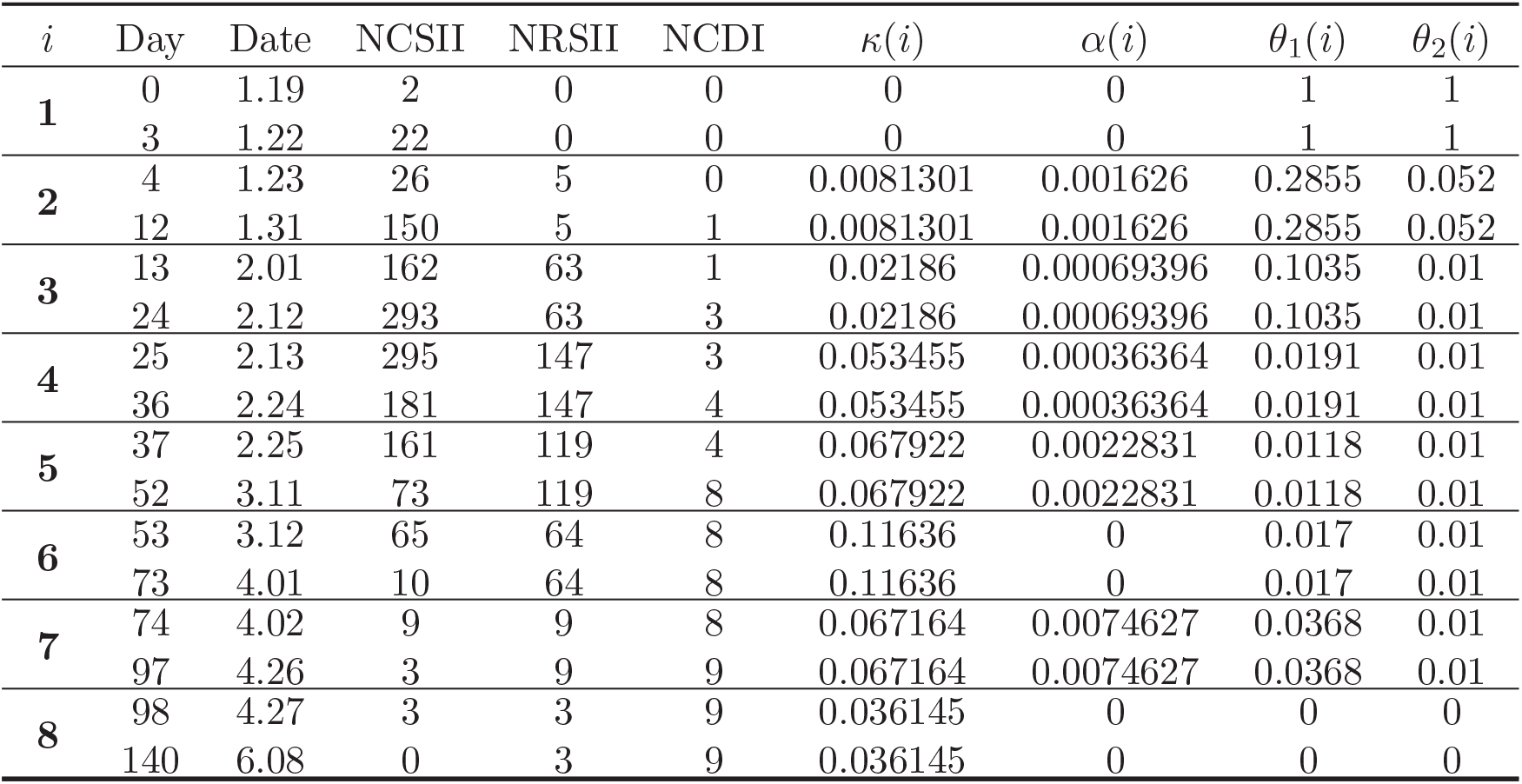
The data of the first COVID-19 epidemic in Beijing on different days and corresponding calculated ; parameters of SARDDE. Where NCSII and NCDI represent the numbers [11] of current symptomatic infected individuals and current died individuals, respectively; NRSII the number [11] of recovered symptomatic infected individuals.

| $i$ | Day | Date | NCSII | NRSII | NCDI | $\kappa(i)$ | $\alpha(i)$ | $\theta_1(i)$ | $\theta_2(i)$ |
| --- | --- | --- | --- | --- | --- | --- | --- | --- | --- |
| **1** | 0 | 1.19 | 2 | 0 | 0 | 0 | 0 | 1 | 1 |
|  | 3 | 1.22 | 22 | 0 | 0 | 0 | 0 | 1 | 1 |
| **2** | 4 | 1.23 | 26 | 5 | 0 | 0.0081301 | 0.001626 | 0.2855 | 0.052 |
|  | 12 | 1.31 | 150 | 5 | 1 | 0.0081301 | 0.001626 | 0.2855 | 0.052 |
| **3** | 13 | 2.01 | 162 | 63 | 1 | 0.02186 | 0.00069396 | 0.1035 | 0.01 |
|  | 24 | 2.12 | 293 | 63 | 3 | 0.02186 | 0.00069396 | 0.1035 | 0.01 |
| **4** | 25 | 2.13 | 295 | 147 | 3 | 0.053455 | 0.00036364 | 0.0191 | 0.01 |
|  | 36 | 2.24 | 181 | 147 | 4 | 0.053455 | 0.00036364 | 0.0191 | 0.01 |
| **5** | 37 | 2.25 | 161 | 119 | 4 | 0.067922 | 0.0022831 | 0.0118 | 0.01 |
|  | 52 | 3.11 | 73 | 119 | 8 | 0.067922 | 0.0022831 | 0.0118 | 0.01 |
| **6** | 53 | 3.12 | 65 | 64 | 8 | 0.11636 | 0 | 0.017 | 0.01 |
|  | 73 | 4.01 | 10 | 64 | 8 | 0.11636 | 0 | 0.017 | 0.01 |
| **7** | 74 | 4.02 | 9 | 9 | 8 | 0.067164 | 0.0074627 | 0.0368 | 0.01 |
|  | 97 | 4.26 | 3 | 9 | 9 | 0.067164 | 0.0074627 | 0.0368 | 0.01 |
| **8** | 98 | 4.27 | 3 | 3 | 9 | 0.036145 | 0 | 0 | 0 |
|  | 140 | 6.08 | 0 | 3 | 9 | 0.036145 | 0 | 0 | 0 |

**Table 2.** The criterions of the asymptotical stability and disease spreading of the disease-free equilibrium of SARDDE at eight time intervals.

| $i$ | Day | $\theta_1(i)$ | $\theta_2(i)$ | $a_{11} + a_{22}$ | $a_{12}a_{21} - a_{11}a_{22}$ | $S_p^1$ | $S_p^2$ |
| --- | --- | --- | --- | --- | --- | --- | --- |
| 1 | 0 | 1 | 1 | 0.69161 | 0.10594 | ∞ | 7.1535 |
| 2 | 4 | 0.2855 | 0.052 | 0.066964 | 0.029394 | 0.4695 | 1.9691 |
| 3 | 13 | 0.1035 | 0.01 | -0.086517 | 0.0079505 | 0.028822 | 1.1134 |
| 4 | 25 | 0.0191 | 0.01 | -0.18146 | -0.0055869 | 0.0032844 | 0.80667 |
| 5 | 37 | 0.0118 | 0.01 | -0.20336 | -0.0087015 | 0.0029741 | 0.62098 |
| 6 | 53 | 0.017 | 0.01 | -0.24559 | -0.014698 | 0.015805 | 0.36595 |
| 7 | 74 | 0.0368 | 0.01 | -0.18892 | -0.0066353 | 0.14924 | 0.62079 |
| 8 | 98 | 0 | 0 | -0.179 | -0.0051636 | 0 | 0 |

Second it needs to determine the parameters 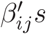 in SARDDE. One can assume that *S* = 1 because the effects of *S* can be deleted by calculated *βij*^′^*s*. This makes the calculated *βij*^′^*s* have general sense. Using the practical data of the first COVID-19 epidemics in Beijing [11](also see the second line in Table 1) selects following initial condition:

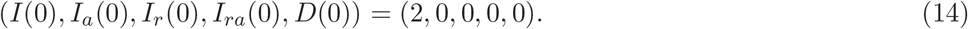

Substitute parameters *κ*(1), *α*(1), *θ*_1_(1) and *θ*_2_(1) listed in Table 1 into system (1). Using a minimization error square criterion:

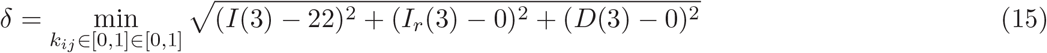

determines 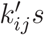.

A group (*β*_11_, *β*_12_, *β*_21_, *β*_22_) that makes *δ* be “smallest” (considering continued simulations) are

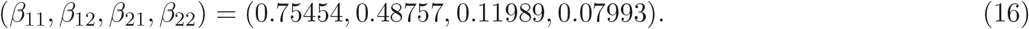

The first 4 days’ simulations of system (1) with the above equation parameters are shown in Figs. 3(a) and 3(b). The simulation results are in good agreement with the reported clinical data (see the first solid and dash lines and legends in Figs.2(a) and (b)).

Third it needs to determine: *θ*_1_(*i*), *θ*_2_(*i*), *i* = 2, 3, *…*, 8. Denote

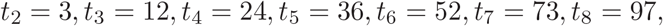

and *D*_*s*_(*t*_*i*_) and *D*_*sr*_(*t*_*i*_) to be the numbers of the Beijing CONVID-19 current symptomatic infected and recovered individuals at *t*_*i*_, respectively, and *D*_*c*_(*t*_*i*_) the cumulative died individuals at *t*_*i*_.

Using the minimization error square criterion:

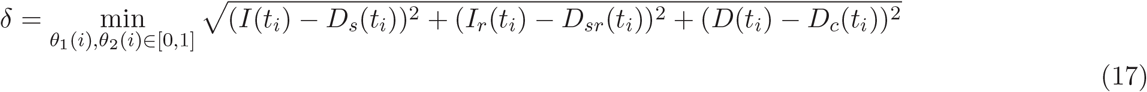

determines the *θ*_1_(*i*) and *θ*_2_(*i*). The calculated results are shown in Table 1.The corresponding simulation results of system (1) are shown in Fig.2(a) and 2(b). Observe that the simulation results of SARDDE model (1) describe well the dynamics of the first COVID-19 epidemic in Beijing.

## Discussions

1. On the days 0th, 3th, 97th, and 140th, the numbers of practical and simulated current asymptomatic individuals are the same. On the days 12th, 36th, and 73th they have only one or two differences. On the days 24th and 52th they have 3 and 5 differences.
2. On the days 0th, 3th, 12th and 97th, the numbers of practical and simulated recovered symptomatic individuals are the same, respectively; On the days 52th, 73th and 140th,, they have 2 or 3 differences, respectively; On the days 24th and 36th,, they have 5 and 7 differences, respectively.
3. The all numbers of practical and simulated cumulative died individuals are the same on the days 0th, 3th, 12th, 24th, 36th, 52th, 73th, 97th and 140th.
4. There is no information on current symptomatic infected and recovered symptomatic infected individuals. But it has reported that after 73th day, April 1, there is no symptomatic infected individuals until the 140th day, June 8[12]. Our simulation results shows that on the 72th day, March 31, the number of the simulated current symptomatic infected individuals was less than one (≈ 0.8), which seems to explain the practical report [11].
5. Computed results (see (16)) of the transmission probabilities *β*_*ij*_ show that the ratio of the probability of asymptomatic and symptomatic individuals infecting susceptible population to become symptomatic individuals is about 0.159 (*β*_21_ : *β*_11_). This suggests that asymptomatic individuals cause lesser symptomatic spread than symptomatic individuals do.
6. The computed results (see (16)) also show that the ratios of the probabilities of asymptomatic and symptomatic individuals infecting susceptible population to become asymptomatic and symptomatic individuals are about 0.646 (*β*_12_ : *β*_11_) and 0.667 (*β*_22_ : *β*_21_), respectively. This suggests that both symptomatic and asymptomatic individuals cause lesser asymptomatic spreads than symptomatic spreads.

(5) The criterions (7) and (8) of the asymptotical stability of the disease-free equilibrium of SARDDE at eight time intervals are listed in the 5th ∼ 8th columns in Table 4. It shows that until the blocking rates (*θ*_1_, *θ*_2_) reach to (0.0191,0.01), the disease-free equilibrium becomes asymptotical stability. The conditions (9) and (10) of disease spreading are listed in the last two columns in Table 4. It shows also that until the blocking rates (*θ*_1_, *θ*_2_) reach to (0.0191, 0.01), the spreading of COVID-19 epidemic can be blocked.

Now assume that after the day 24th, February 12, it still keeps the blocking rates (*θ*_1_(3), *θ*_2_(3)), the cure rates (*κ*(3), *κ*_*a*_(3)), and the died rate *α*(3) until the day 140th, June 8. The simulation results of SARDDE are shown in Figs 4(a) and 4(b). Observe that the numbers of the current symptomatic and asymptomatic infected individuals reach to about 1.899e5 and 4678, respectively. The numbers of cumulative recovered symptomatic and died individuals reach to about 11983 and 2358, respectively.

**Figure 4:**
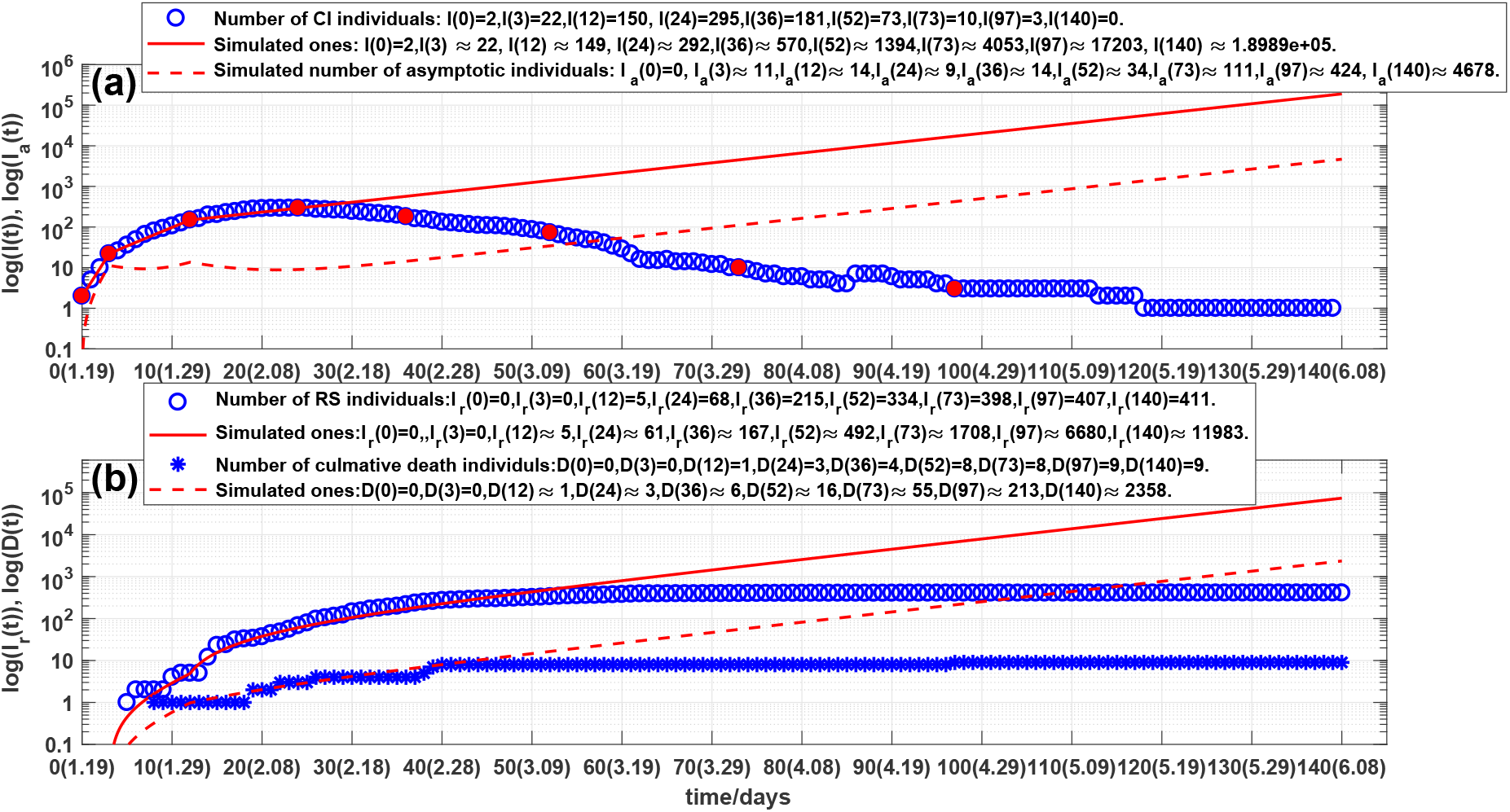
Outcomes of the numbers of:(a) current symptomatic individuals, representing by circles. Solid line and dash line are the stimulated current symptomatic and asymptomatic individuals of system (1); (b) cumulative recovered symptomatic and died individuals, representing by circles and stars, respectively. Solid line and dash line are corresponding simulations of system (1).

Furthermore assume that after the day 36th, February 24, it still keeps the blocking rates (*θ*_1_(4), *θ*_2_(4)), the cure rates (*κ*(4), *κ*_*a*_(4)), and the died rate *α*(4) until the day 140th, June 8. The simulation results of SARDDE are shown in Figs 5(a) and 5(b). Observe that the numbers of the current symptomatic and asymptomatic infected individuals are about three and less than one, respectively; The numbers of cumulative recovered symptomatic and died individuals are about 453 and 5, respectively. The results suggest that using the data before the day 37th (about two weeks after the turning point) is able approximately to estimate the following outcome of the the first COVID-19 academic in Beijing.

**Figure 5:**
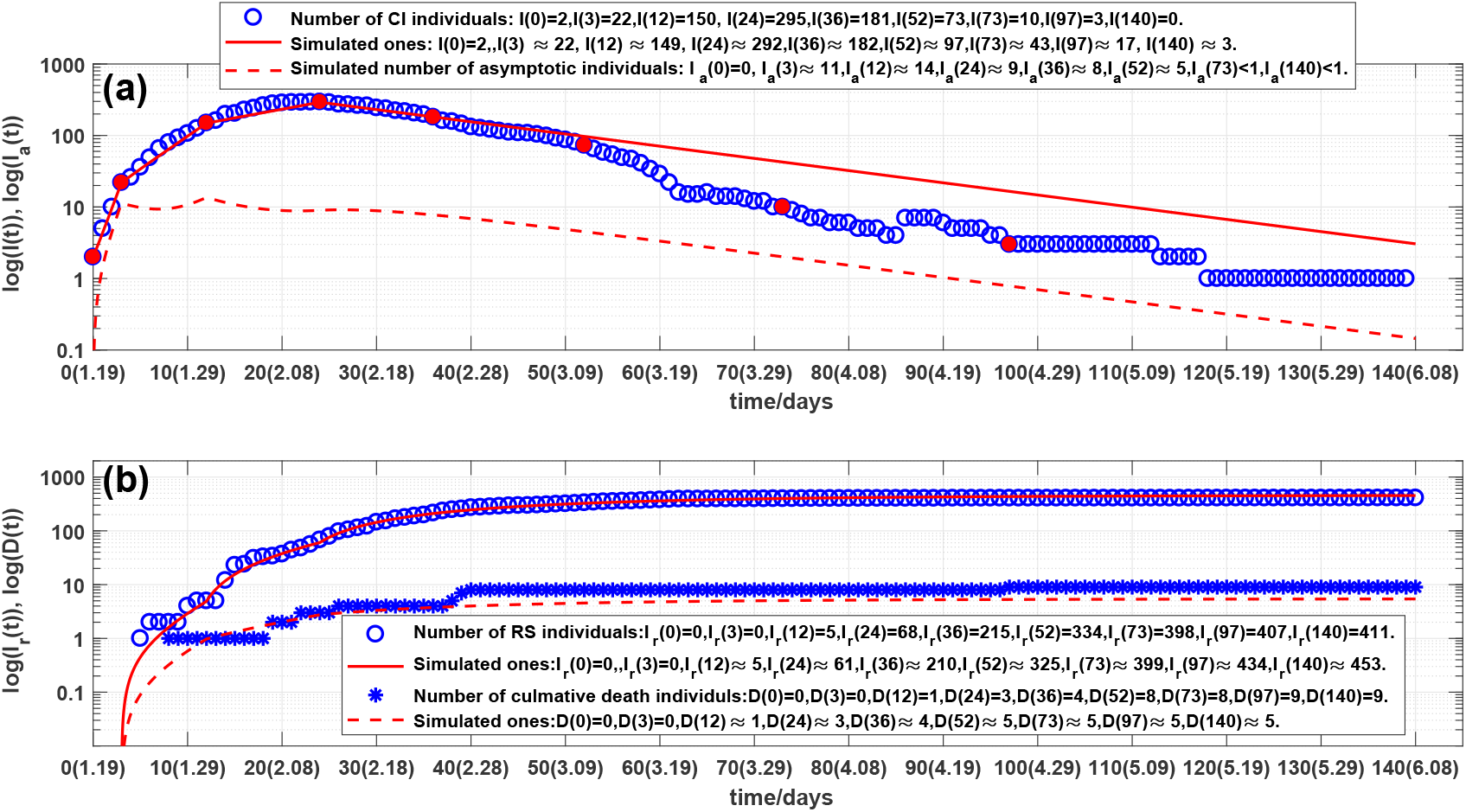
Outcomes of the numbers of:(a) current symptomatic individuals, representing by circles. Solid line and dash line are the stimulated current symptomatic and asymptomatic of system (1); (b) cumulative recovered symptomatic and died individuals, representing by circles and stars, respectively. Solid line and dash line are corresponding simulations of system (1).

In summary, SARDDE model (1) can simulate the outcomes of the first COVID-19 epidemic in. Beijing. The calculated equation parameters can help us to understand and explain the mechanism of epidemic diseases and control strategies for the event of the practical epidemic.

### 3.2 Simulation and prediction of the second COVID-19 epidemic in Beijing

Until June 10, 2020, the Beijing whole city continual 56 days has no new reports of the locally 45 confirmed COVID-19 cases. There have been 11 districts in all 15 districts which continually have no reported locally COVID-19 case over 100 days. However in June 11, Xinfadi in Tongzou district appeared a COVID-19 confirmed case. Thus has caused the second wave COVID-19 epidemic in Beijing.

A total of 335 locally diagnosed cases were reported during the 2th wave COVID-19 epidemics. After 56 days, all COVID-19 patients were cured. The medical personnel has realized the zero infection. This event of Xinfadi COVID-19 epidemic provides a valuable example of accurate preventing and controlling strategies and excellent clinical treatments.

#### 3.2.1 Model

Similar to Section 2.1 The transition among these states is governed by the following rules (Flowchart of the rules is shown in Fig.6, where *S* represents susceptible population.)

**Figure 6:**
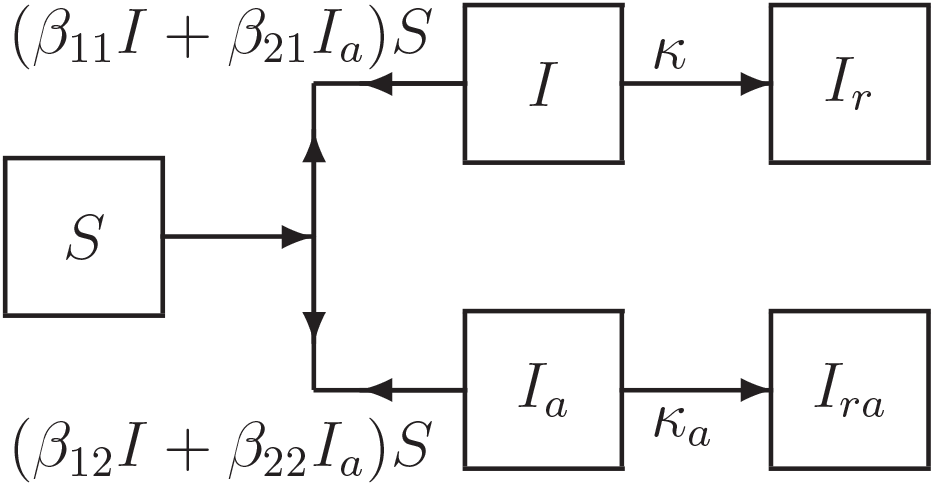
Flowchart of disease transmission among susceptible population S, current symptomatic infected individuals I, current asymptomatic but infected individuals *I*_*a*_ recovered symptomatic infected individuals *I*_*r*_, recovered asymptomatic but infected individuals *I*_*ra*_.

Similar to Section 2.1, assume that the dynamics of an epidemic can be described by *m* time intervals. At *i*th interval, the model has the form:

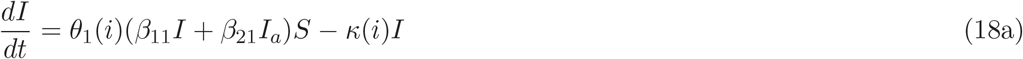

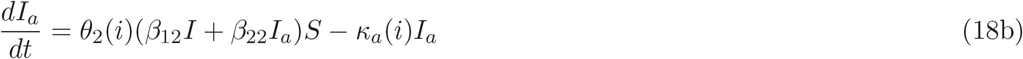

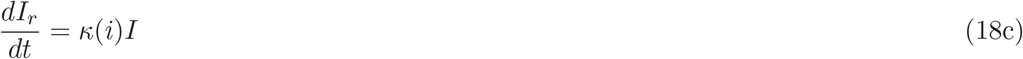

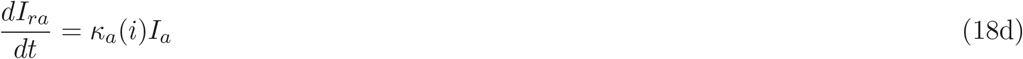

Then system (18) has a disease-free equilibrium:

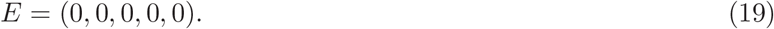

#### 3.2.2 Stability of disease-free equilibrium

The stability of system (18) is determined by the first two equations (18a) and (18b). Denote in (18a) and (18b):

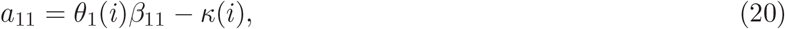

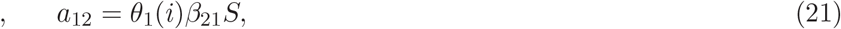

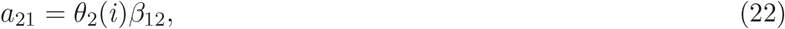

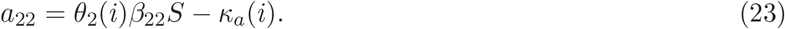

Then at the disease-free equilibrium of system(18), the Jacobian matrix of (18a) and (18b) is

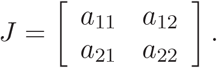

Solving the corresponding eigenequation obtains 2 eigenvalues:

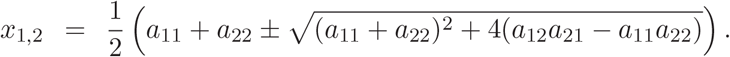

Therefore it obtains the following:

##### Theorem 3

*Suppose that a*_11_, *a*_12_, *a*_21_ *and a*_22_ *are defined by (20)-(23) then the disease-free equilibrium E of system (1) is globally asymptotically stable if, and only if, the following inequalities hold:*

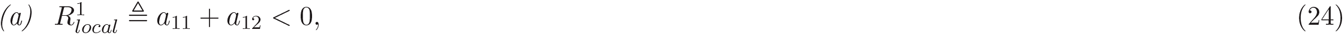

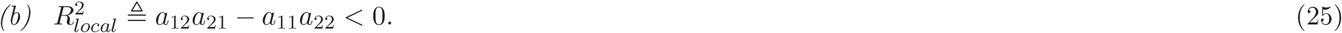

#### 3.2.3 The necessary condition of disease spreading

If an epidemic can occur, then

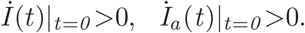

This implies that

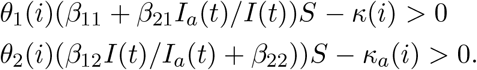

Solving the above inequalities gives the following

##### Theorem 4

*If system (1) satisfy the following inequalities*

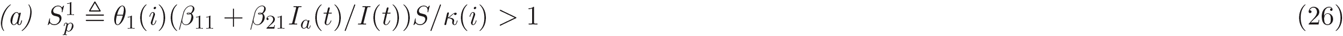

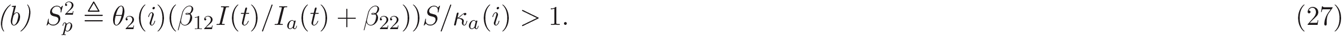

*then a disease transmission will occur*.

#### 3.2.4 Simulations

Based on the reported clinical COVID-19 epidemic data from June 11 to August 6, 2020 in Beijing [11], this Section will discuss the applications of above theoretical results. The numbers of current symptomatic infected individuals, and current asymptomatic but infected individuals are showed in Fig, 7(a) by circles and diamonds, respectively. The numbers of current recovered symptomatic infected individuals, and current recovered asymptomatic but infected individuals are showed in Fig. 7(b) by circles and diamonds, respectively.

**Figure 7:**
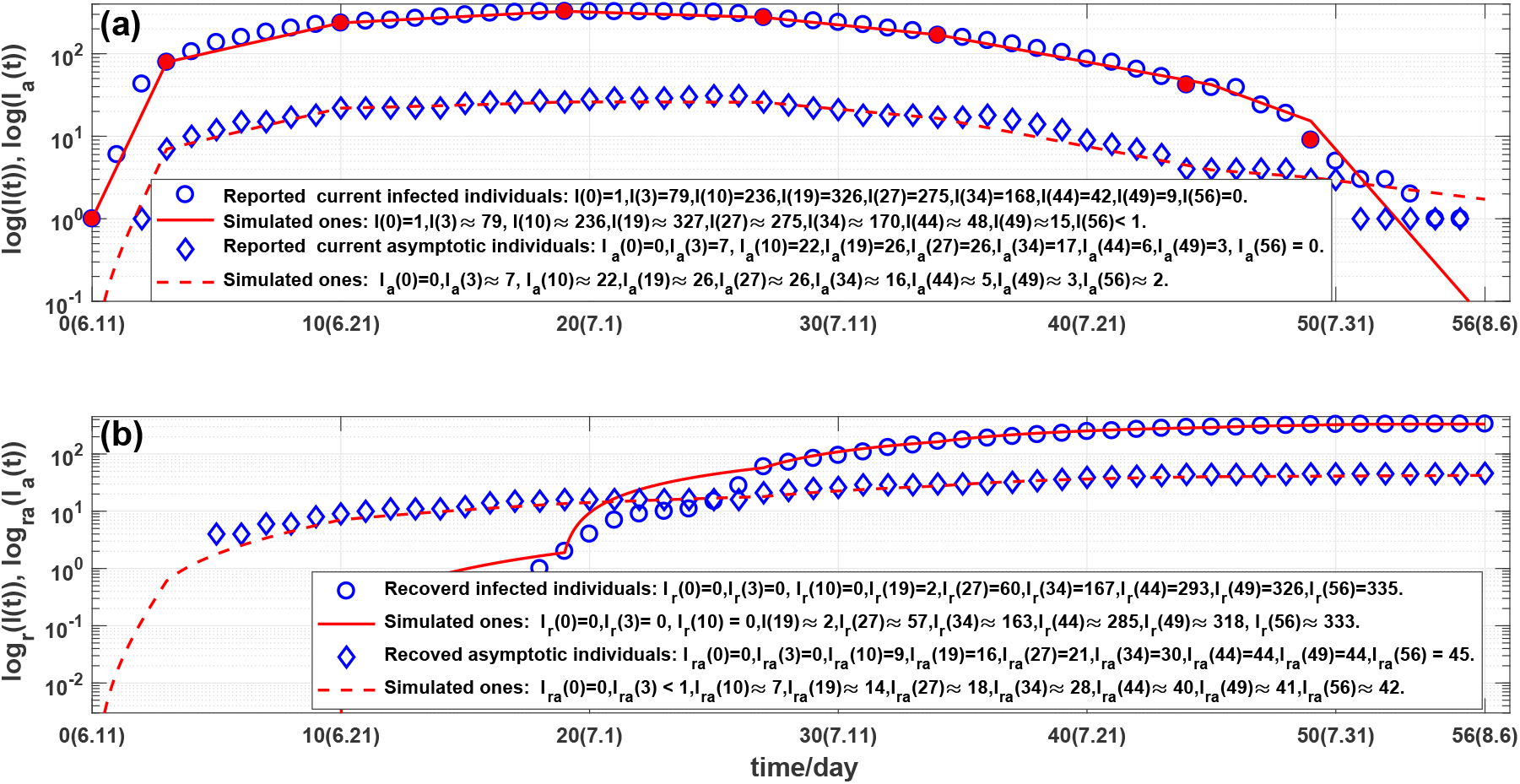
Outcomes of the numbers of:(a) current symptomatic individuals, representing by circles. Solid line and dash line are the stimulated current symptomatic and asymptomatic individuals of system (18); (b) cumulative recovered symptomatic and died individuals, representing by circles and stars, respectively. Solid line and dash line are corresponding simulations of system (18).

The number of current infected individuals was risen rapidly in the first 4 days (see Fig. 7(a)). The number of current infected individuals reached the highest 326 on the day 19th, June 30 (see Fig. 7(a)), and then after the day 27th, July 7, declined rapidly. The corresponding cumulative number of recovered symptomatic and asymptomatic individuals has risen rapidly after the day 27th, July 7 (see Fig. 7(b)). Observe from the Figs. 7(a) and 7(b) that the overall changes in the number of current firmed infections are not subject to the law of exponential changes, but the data can be approximated in good agreement with 8 straight lines in log scale (see Fig. 7). This phenomenon can be explained as: different medical measures and prevention and control strategies have been adopted at the different 8 time intervals. Therefore the *i* in model (18) satisfies

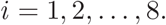

First it needs to determine the parameters *κ*(*i*), *κ*_*a*_(*i*). Denote *s*_1_(*i*) and *s*_2_(*i*) to be the days that the old patients and the new patients stayed in the hospital during *i*th time interval. Denote *R*(*i*) and *R*_*a*_(*i*) to be the numbers of the recovered symptomatic patients and asymptomatic patients during *i*th time interval, respectively. Then *κ*(*i*) and *κ*_*a*_(*i*) can be defined by

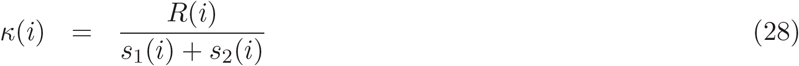

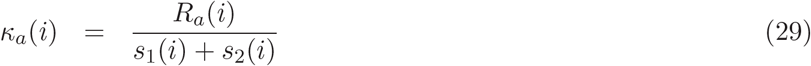

The calculated *κ*(*i*)^′^*s* and *κ*_*a*_(*i*)^′^*s* are shown in Table 3, in which the data of *κ*_*a*_(1) have used the first five day’s data because there are 4 asymptomatic but infected patients who had recovered on the 5th day (see Fig7. (b) and Table 3).

**Table 3.** The data of Xingodi COVID-19 epidemics on 18 different days and corresponding calculated parameters of system (18). Where NCSII and NCAII represent the numbers of current symptomatic infected individuals and current asymptomatic infected individuals, respectively; NRSII and NRAII represent the numbers of recovered symptomatic infected individuals and recovered asymptomatic but infected individuals, respectively.

| $i$ | Day | Date | NCSII | NCAII | NRSII | NRAII | $\kappa(i)$ | $\kappa_a(i)$ | $\theta_1(i)$ | $\theta_2(i)$ |
| --- | --- | --- | --- | --- | --- | --- | --- | --- | --- | --- |
| **1** | 0 | 6.11 | 1 | 0 | 0 | 0 | 0 | 0.13333 | 1 | 1 |
|  | 1 | 6.12 | 6 | 0 | 0 | 0 | 0 | 0.13333 | 1 | 1 |
|  | 2 | 6.13 | 43 | 1 | 0 | 0 | 0 | 0.13333 | 1 | 1 |
|  | 3 | 6.14 | 79 | 7 | 0 | 0 | 0 | 0.13333 | 1 | 1 |
| **2** | 4 | 6.15 | 106 | 10 | 0 | 0 | 0 | 0.069231 | 0.1075 | 0.1497 |
|  | 5 | 6.16 | 137 | 12 | 0 | 4 | 0 | 0.069231 | 0.1075 | 0.1497 |
|  | 10 | 6.21 | 236 | 22 | 0 | 9 | 0 | 0.069231 | 0.1075 | 0.1497 |
| **3** | 11 | 6.22 | 249 | 22 | 0 | 10 | 0.000728 | 0.030837 | 0.0250 | 0.0298 |
|  | 19 | 6.30 | 326 | 26 | 2 | 16 | 0.000728 | 0.030837 | 0.0250 | 0.0298 |
| **4** | 20 | 7.01 | 325 | 28 | 4 | 16 | 0.022925 | 0.020080 | 0.0012 | 0.0123 |
|  | 27 | 7.08 | 275 | 26 | 60 | 21 | 0.022925 | 0.020080 | 0.0012 | 0.0123 |
| **5** | 28 | 7.09 | 263 | 24 | 72 | 23 | 0.069301 | 0.065217 | 0 | 0.001 |
|  | 34 | 7.15 | 168 | 17 | 167 | 30 | 0.069301 | 0.065217 | 0 | 0.001 |
| **6** | 35 | 7.16 | 158 | 17 | 177 | 30 | 0.12647 | 0.12174 | 0 | 0 |
|  | 44 | 7.25 | 42 | 4 | 293 | 44 | 0.12647 | 0.12174 | 0 | 0 |
| **7** | 45 | 7.26 | 39 | 4 | 296 | 44 | 0.25385 | 0.052632 | 0 | 0 |
|  | 49 | 7.30 | 9 | 3 | 326 | 44 | 0.25385 | 0.052632 | 0 | 0 |
| **8** | 50 | 7.31 | 5 | 3 | 330 | 44 | 0.79167 | 0.086957 | 0 | 0 |
|  | 56 | 8.06 | 0 | 0 | 335 | 45 | 0.79167 | 0.086957 | 0 | 0 |

Second it needs to determine the parameters 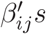 in system (18). One can assume that *S* = 1 because the effects of S can be deleted by calculated 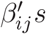. This makes the calculated 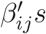 have general sense. Using the practical data of Xinfadi COVID-19 epidemic (see the second line in Table 3) selects following initial condition

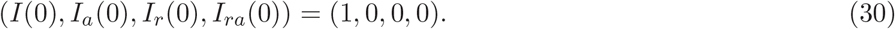

Substitute parameters *κ*(1), *κ*_*a*_(1), *θ*_1_(1) and *θ*_2_(1) listed in Table 3 into system (18). Using a minimization error square criterion:

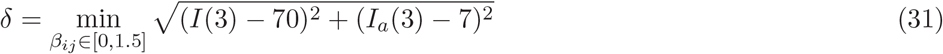

determines 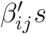. A group 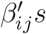 that makes *δ* be “smallest” (considering continued simulations) are

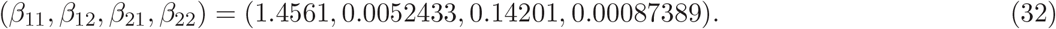

The simulations of system (18) with above equation parameters are shown in Figs. 7(a) and 7(b). Observe that the simulation results are in good agreement with the reported first 4 days’ clinical data (see the solid and dash lines in Figs. 7(a) and 7(b)).

Third it needs to determine: *θ*_1_(*i*), *θ*_2_(*i*), *i* = 2, 3, *…*, 8. Denote

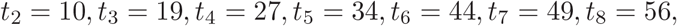

and *D*_*s*_(*t*_*i*_) and *D*_*sr*_(*t*_*i*_) to be the numbers of the current symptomatic infected and recovered individuals at *t*_*i*_, respectively, and *D*_*c*_(*t*_*i*_) the cumulative died individuals at *t*_*i*_.

Using the minimization error square criterion:

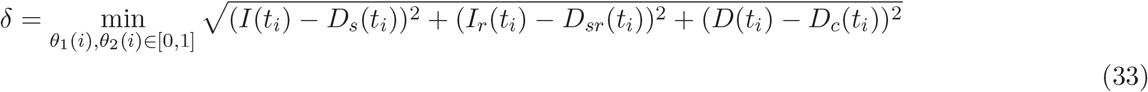

determines the *θ*_1_(*i*) and *θ*_2_(*i*). The calculated results are shown in Table 3.The corresponding simulation results of system (18) are shown in Fig.7(a) and 7(b). Observe that the simulation results of model (18) describe well the dynamics of the second COVID-19 epidemic in Beijing.

## Discussions

1. On the days 0th, 3th,, 10th, 27th, and 56th, the numbers of the practical and simulated current symptomatic individuals are the same. On the days 19 and 34th,, they have only one and two differences; On the days 44th and 49th, they have six differences.
2. On the days 0th, 3th, 10th, 19th, 27th, 49th and 56th, the numbers of the practical and simulated current asymptotic individuals are the same; On the days 34th and 44th, they have only one difference.
3. On the days 0th, 3th,, 10th, and 19th, the numbers of practical and simulated recovered symptomatic individuals are the same, respectively; On the days 27th and 56th, they have only 3 and 2 differences, respectively; On the days 34th it has 4 differences; On the days 44th and 49th, they have 8 differences.
4. On the days 0th and 3th,, the numbers of practical and simulated recovered asymptomatic individuals are the same, respectively; On the days 10th 19th and 34th, they have only 2 differences; On the days 27th, 49th and 56th, they have three differences, respectively; On the day 44th, it has 4 differences.
5. Computed results (see (32)) show that the ratio of the probability of asymptomatic and symptomatic individuals infecting susceptible population to become symptomatic individuals is about 0.36% (*β*_21_: *β*_11_). This suggests that asymptomatic individuals cause lesser symptomatic spread than symptomatic individuals do.
6. Computed results (see (32)) also show that the ratios of the probabilities of asymptomatic and symptomatic individuals infecting susceptible population to become asymptomatic and symptomatic individuals are about 9.75% (*β*_12_ : *β*_11_) and 16.67% (*β*_22_ : *β*_21_), respectively. This suggests that both symptomatic and asymptomatic individuals cause lesser asymptomatic spread than symptomatic spread.
7. The criterions of the stability of the disease-free equilibrium of system (18) at five time intervals are listed in Table 4. It shows that until the blocking rates (*θ*_1_, *θ*_2_) reach to (0.0012, 0.0123), the disease-free equilibrium becomes asymptotical stable.

Now assume that it keeps still the blocking rates (*θ*_1_(3), *θ*_2_(3)) and the cure rates (*κ*(3), *κ*_*a*_(3)) until the day 56th, August 6. The simulation results of system (18) are shown in Fig. 8. Observe that the numbers of the current symptomatic and asymptomatic infected individuals reach to 1235 and 80, respectively; The numbers of cumulative recovered symptomatic and asymptomatic infected individuals reach to 21 and 66, respectively

**Table 4.** The criterions of the asymptotical stability and disease spreading of the disease-free equilibrium of system (18) at eight time intervals.

| $i$ | Day | $\theta_1(i)$ | $\theta_2(i)$ | $a_{11} + a_{22}$ | $a_{12}a_{21} - a_{11}a_{22}$ | $S_p^1$ | $S_p^2$ |
| --- | --- | --- | --- | --- | --- | --- | --- |
| 1 | 3 | 1 | 1 | 1.4613 | 0.19361 | $+\infty$ | 12.02 |
| 2 | 10 | 0.1075 | 0.1497 | 0.15709 | 0.010998 | $+\infty$ | 3.2635 |
| 3 | 19 | 0.0252 | 0.0298 | 0.036071 | 0.0010982 | 0.1627 | 1.7398 |
| 4 | 27 | 0.0012 | 0.0123 | -0.021363 | -0.00045639 | 0.00029997 | 0.8738 |
| 5 | 34 | 0 | 0.001 | -0.069301 | -0.0045195 | 0 | 0.022388 |
| 6 | 44 | 0 | 0 | -0.12647 | -0.016547 | 0 | 0 |
| 7 | 49 | 0 | 0 | -0.25385 | -0.013361 | 0 | 0 |
| 8 | 56 | 0 | 0 | -0.79167 | -0.068841 | 0 | 0 |

**Figure 8:**
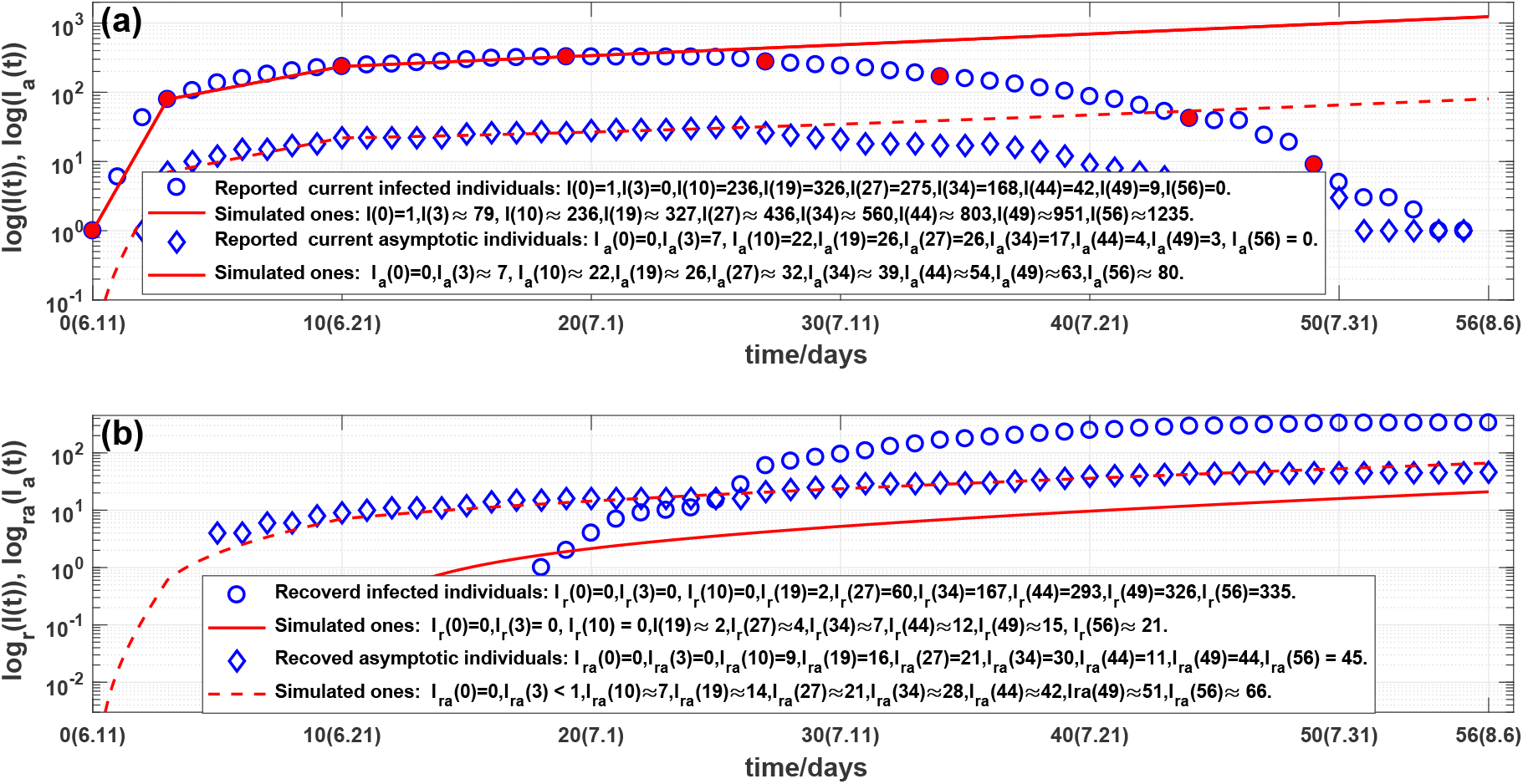
Outcomes of the numbers of:(a) current symptomatic and asymptomatic individuals, representing by circles and diamonds. Solid line and dash line are the corresponding stimulated ones of system (18); (b) cumulative recovered symptomatic and asymptomatic individuals, representing by circles and diamonds. Solid line and dash line are the corresponding stimulated ones of system (18).

Furthermore assume that after the day 34th, July 14, it still keeps the blocking rates (*θ*_1_(5), *θ*_2_(5)), the cure rates (*κ*(5), *κ*_*a*_(5)) until the day 56th, August 6. The simulation results of system (18) are shown in Figs. 9(a) and 9(b). Observe that the numbers of the current symptomatic and asymptomatic infected individuals are about 37 and 4, respectively. The numbers of cumulative recovered symptomatic and asymptomatic individuals are about 296 and 40, respectively. The results suggest that using the data before the day 34th (about two weeks after the turning point) is able approximately to estimate the following outcome of the the second COVID-19 academic in Beijing.

**Figure 9:**
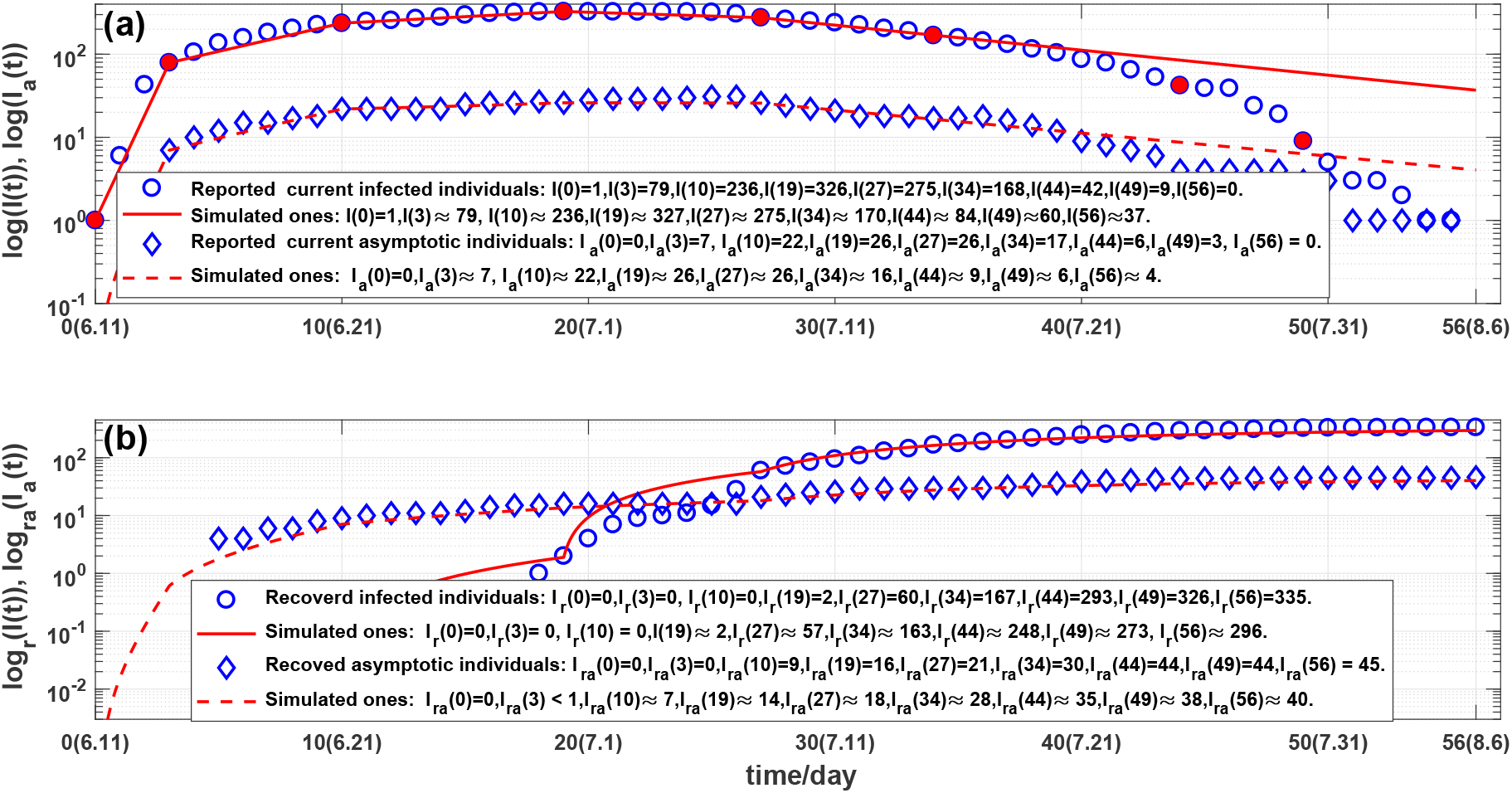
Outcomes of the numbers of:(a) current symptomatic and asymptomatic individuals, representing by circles and diamonds. Solid line and dash line are the corresponding stimulated ones of system (18); (b) cumulative recovered symptomatic and asymptomatic individuals, representing by circles and diamonds. Solid line and dash line are the corresponding stimulated ones of system (18).

## 4 Conclusions

The main contributions of this paper are summarized as follows:

1. Proposed SARDDE model with five states: current symptomatic and asymptomatic infected individuals, cumulative recovered symptomatic and asymptomatic infected individuals, died individuals.
2. Provided the criterion inequalities for the asymptotical stability of the disease free equilibrium point of SARDDE (see Theorem 1 and Theorem 3).
3. Given the criterion inequalities for epidemic transmission (see Theorem 2 and Theorem 4) of the symptomatic and asymptomatic infections.
4. Used the reported data of the first COVID19 epidemic in Beijing [11] to determine the parameters in SARDDE, and carry out simulations. The simulation results are in good agreement with the reported clinic data [11].
5. In systems (1) and (18), assume, respectively, that after the day 24th and the day 19th if still keeps the blocking rates (*θ*_1_(3), *θ*_2_(3)), the cure rates (*κ*(3), *κ*_*a*_(3)) and the died rate *α*(3) until the day 140th, June 8 and the day 56, August 6. Virtual simulations of systems (1) and (18) suggest that even the a blocking rate to symptomatic individuals reaches to about 90%, the COVID-19 epidemic can still spread and reach very height levels (see Figs. 4 and 7). Therefore the strict prevention and control strategies implemented by Beijing government is not only very effective but also completely necessary.
6. Simulations showed that using the data form the beginning to the day after about two weeks the turning points, we can estimate well or approximately the following outcomes of the first or second COVID-19 academics in Beijing.

9 In the case of the first COVID-19 academic in Beijing, the proposed model SARDDE is simpler that the previous ones [7, 8], and it is only used one assumption (13). However it seems to be able better to describe and explain the practical data. [11] although the lack of symptomatic infected individuals.
10 In the case of the second COVID-19 academic in Beijing, the proposed model (18) describe well the practical data [11].

It is expected that researches can provide better understanding, explaining, and dominating for epidemic spreads, preventions and controls.

## Data Availability

All reported data cited in the manucsript were selected from
the web site of Health Commision of Beijing (http://wjw.beijing.gov.cn/ see Ref.[11]).
Now I have found that somme earler data are not availble. However I have downloaded
all related web pages.

## Funding

The author declares no potential conflict of interest.

## Conflict of Interest

The author declares no potential conflict of interest.

## Note

The previous versions of the manuscript have been published online [12, 13].

## Footnotes

1 In the cases that some reported data crossed one day, we assign approximately numbers according to the ratios of time intervals.

## Notes

### Competing Interest Statement

The authors have declared no competing interest.

### Author Declarations

No any IRB/oversight body that provided approval or exemption for the research described.

